# Ultrastructural characterisation of young and aged dental enamel by atomic force microscopy

**DOI:** 10.1101/2022.03.09.22271977

**Authors:** Camila Leiva-Sabadini, Christina MAP Schuh, Nelson P Barrera, Sebastian Aguayo

## Abstract

Recent advances in atomic force microscopy (AFM) have allowed the characterisation of dental-associated biomaterials and biological surfaces with high-resolution and minimal sample preparation. In this context, the topography of dental enamel – the hardest mineralised tissue in the body – has been explored with AFM-based approaches at the micro-scale. With age, teeth are known to suffer changes that can impact their structural stability and function; however, changes in enamel structure because of ageing have not yet been explored with nanoscale resolution. Therefore, the aim of this exploratory work was to optimise an approach to characterise the ultrastructure of dental enamel and determine potential differences in topography, hydroxyapatite (HA) crystal size, and surface roughness at the nanoscale associated to ageing. For this, a total of six teeth were collected from human donors from which enamel specimens were prepared. By employing AC mode imaging, HA crystals were characterised in both transversal and longitudinal orientation with high-resolution in environmental conditions. Sound superficial enamel displayed the presence of a pellicle-like coating on its surface, that was not observable on cleaned specimens. Acid-etching exposed crystals that were imaged and morphologically characterised in highresolution at the nanoscale in both the external and internal regions of enamel in older and younger specimens. Our results demonstrated important individual variations in HA crystal width and roughness parameters across the analysed specimens; however, an increase in surface roughness and decrease in HA width was observed for the pooled older external enamel group compared to younger specimens. Overall, high-resolution AFM was an effective approach for the qualitative and quantitative characterisation of human dental enamel ultrastructure at the nanometre range. Future work should focus on exploring the ageing of dental enamel with increased sample sizes to compensate for individual differences as well as other potential confounding factors such as behavioural habits and mechanical forces.

## 1. Introduction

Human teeth are complex organs located in the oral cavity, made up by three mineralised tissues (enamel, dentin, cementum) and one non-mineralised tissue (dental pulp). Amongst these, dental enamel is the outermost mineralised layer that coats the visible crown portion of the tooth^1,2^. As such, it is in direct contact with the oral environment and thus is a crucial substrate for the chemical and mechanical protection of underlying tissues such as dentin and the dental pulp. It is mostly constituted by a mineral phase of hydroxyapatite (HA) – a form of calcium apatite - although there are also very minor amounts of specialised enamel proteins such as amelogenins and enamelins that act as scaffolding for mineralisation^3^. This high degree of mineralisation makes dental enamel the hardest tissue in the human body and grants it mechanical stability to endure cyclic force loading during repetitive mastication. Protein-protein and protein-mineral interactions regulate the morphology and three-dimensional orientation of hydroxyapatite crystals and determine the mechanical properties of enamel such as high surface hardness and resistance to fracture^4,5^.

Within enamel, HA crystals are known to be ∼50-70 nm wide and up to several microns in length^6,7^, and can be found aligned within anatomical structures known as enamel rods that enhance the mechanical properties of the tissue^8^. Besides being an important structural tissue, dental enamel is also an important substrate for the adhesion of biomaterials during restorative treatments. In recent years, the development of adhesive dentistry procedures allows the direct bonding of composite resin materials to tooth structures including enamel^9^. These techniques involve an initial demineralisation of the enamel surface with phosphoric acid to controllably remove HA crystals. This leads to an increase in surface energy and microroughness and facilitates the formation of a strong restorative-tooth interphase that will increase the long-term success of treatment. As dental restorations can be placed either on superficial enamel or deeper within the tissue, the ultrastructure of enamel at different depths is clinically relevant and plays an important role in adhesive dentistry.

Currently, it is known that dental tissues can suffer important mechanical and biological changes with ageing. Some relevant ageing markers in teeth include dental pulp fibrosis^10^, dentinal sclerosis, and increased elasticity, as well as age-dependent accumulation of glycation end-products^11^. In enamel, age-associated alterations are mostly a result of cyclic exposure to mechanical forces (such as mastication) and erosion (due to dietary or intrinsic factors) that modify the structure and roughness of enamel over time9. Despite these observations, little is known regarding ageing-associated ultrastructural changes in enamel at the nanoscale that could potentially impact tissue mechanics, resistance to wear, and integration with restorative biomaterials.

In recent years, the use of atomic force microscopy (AFM) has allowed researchers to characterise biological substrates and tissues with nanoscale resolution^12,13^. For AFM, samples require minimal preparation and can be observed in near-physiological conditions, resulting in huge advantages for the exploration of human tissue samples and specimens^14^. Furthermore, AFM is not limited to topographical characterisation as it can also provide important quantitative information such as surface roughness and mechanical properties of specimens and substrates^15^. Previous work has utilised AFM to examine the microstructure of enamel surfaces after acid etching but mostly focused on observing changes at the microscale range^16–18^; however, potential enamel ultrastructural changes associated to tooth ageing have not yet been explored with these techniques.

Thus, the aim of this exploratory work was to optimise an approach to characterise the ultrastructure of dental enamel with nanoscale resolution in both younger and older tooth specimens, in order to determine potential differences in topography, HA crystal size, and surface roughness at the nanoscale. Overall, understanding potential changes in enamel ultrastructure between younger and older tooth specimens could potentially aid in the development of novel therapeutic approaches regarding adhesive dentistry focused on the elderly.

## 2. Materials and methods

### 2.1. Sample collection

For enamel specimen collection, a total of 6 permanent teeth extracted due to orthodontic or restorative treatment were obtained following informed consent (local ethical approval #180426002). All tooth samples presented sound enamel at the moment of extraction with the absence of restorations or active caries. Teeth were initially washed and stored in 70% ethanol solution for 72 hours for decontamination, washed 3x in PBS, and air-dried.

### 2.2. Specimen preparation

Following sample collection, teeth were embedded in acrylic resin (Marche Acrylics, Chile) and sectioned transversally with an SP1600 hard tissue microtome (Leica Biosystems, US) to obtain 200 µm specimens for each tooth. After sectioning, tooth samples were treated with a 37% phosphoric acid solution (Sigma-Aldrich, US) for 30 sec, washed 3x with ultrapure water (dH_2_O), and air-dried. Furthermore, non-sectioned enamel specimens, corresponding to non-occlusal regions for each tooth, were also collected to directly visualise the external surface of enamel.

Initial specimen characterisation was carried out with light microscopy and scanning electron microscopy (SEM). Briefly, specimens were immobilised onto glass cover slides and visualised with a Panthera U (Motic, China) light microscope, obtaining images at 4x and 10x magnification. Subsequently, selected specimens were submerged in a solution of 2.5% glutaraldehyde for 24 hours, dehydrated with an increasing 25%, 50%, 70%, 90%, 100% ethanol series, gold sputter-coated, and imaged with a Hitachi TM3000 SEM (Tokyo, Japan) with an acceleration voltage of 5 kV.

### 2.3. Atomic force microscopy (AFM)

All AFM experiments were performed with an Asylum MFP 3D-SA AFM (Asylum Research, US) in intermittent contact (AC mode) under environmental conditions, utilising SCOUT 350 RAu probes (NuNano, Bristol, United Kingdom). Briefly, superficial and sectioned enamel specimens were cleaned with a stream of N_2_ air and immobilised onto metal discs with the aid of double-sided tape. Initial exploratory 10×10 µm scans were performed on each sample, from where 1×1 µm representative images were obtained for nanoscale visualisation. Height, amplitude, and phase channels were recorded, and gain parameters were adjusted in real time to optimise image acquisition at 256×256 pixels. For sectioned specimens, images were acquired at both external (adjacent to the enamel surface) and internal (adjacent to the dentin-enamel junction) regions of enamel for each specimen. A total of 3 representative images were acquired for each specimen at each location.

### 2.4. Data and statistical analysis

All AFM images were processed with the Gwyddion 2.6 software. Selected trace profiles and 3D images were obtained from the height channel. HA crystal sizes were measured and determined with ImageJ Fiji v2.3.0 software and graphed as violin plots (median and quartiles) in Graphpad Prism 9. RMS roughness was determined in Gwyddion across three independent 1×1 µm scans per specimen and expressed as mean ± standard deviation. Normality tests and statistical analysis were performed in Graphpad Prism 9 software with either one-way ANOVA with Tukey’s multiple comparisons, or Kruskal-Wallis test with Dunn’s multiple comparison tests, considering significance at p values <0.05.

## 3. Results and Discussion

### 3.1. Characterisation of superficial human enamel

For this exploratory study, we obtained dental enamel specimens from a total of 6 donors from two different age groups. All employed teeth were free of dental caries lesions, evident chemical modifications, or previous restorative procedures. Three of the samples were obtained from younger individuals (20-26 years old), whereas the other three samples were collected from older individuals (50-69 years old). For all cases, an appropriate amount of tooth slices (specimens) was obtained via hard tissue microtome sectioning, and sound enamel was also obtained for superficial sample characterisation.

Enamel is the most external tissue covering the surface of the tooth, and as such, it lies in direct contact with the oral microenvironment^19^. Thus, as an initial step, the visualisation of human enamel with AC mode AFM was done directly on the tooth surface before any cleaning procedure. As such, the presence of a pellicle-like structure was observed directly on top of the surface of enamel (**Figures 1A and 1B**). This can be better observed with the phase channel that highlights the differences in physicochemical properties between the pellicle and parts of the underlying enamel surface that are visible (**Figures 1C and 1D**). It is known that within the oral cavity, enamel is coated with a pellicle consisting of salivary proteins and other macromolecules such as amylase, mucin, and lipids, amongst others^20^. Therefore, the presence of a pellicle-like structure on the surface of extracted human tooth enamel is expected during the direct examination under AFM with no previous sample preparation or cleaning. It remains likely that the pellicle observed in the present samples may be due to minor remnants of the salivary pellicle that survived sample preparation, as previous work has shown that intact salivary pellicles can form clusters up to 80nm height when observed with AFM^21^.

**Figure 1:**
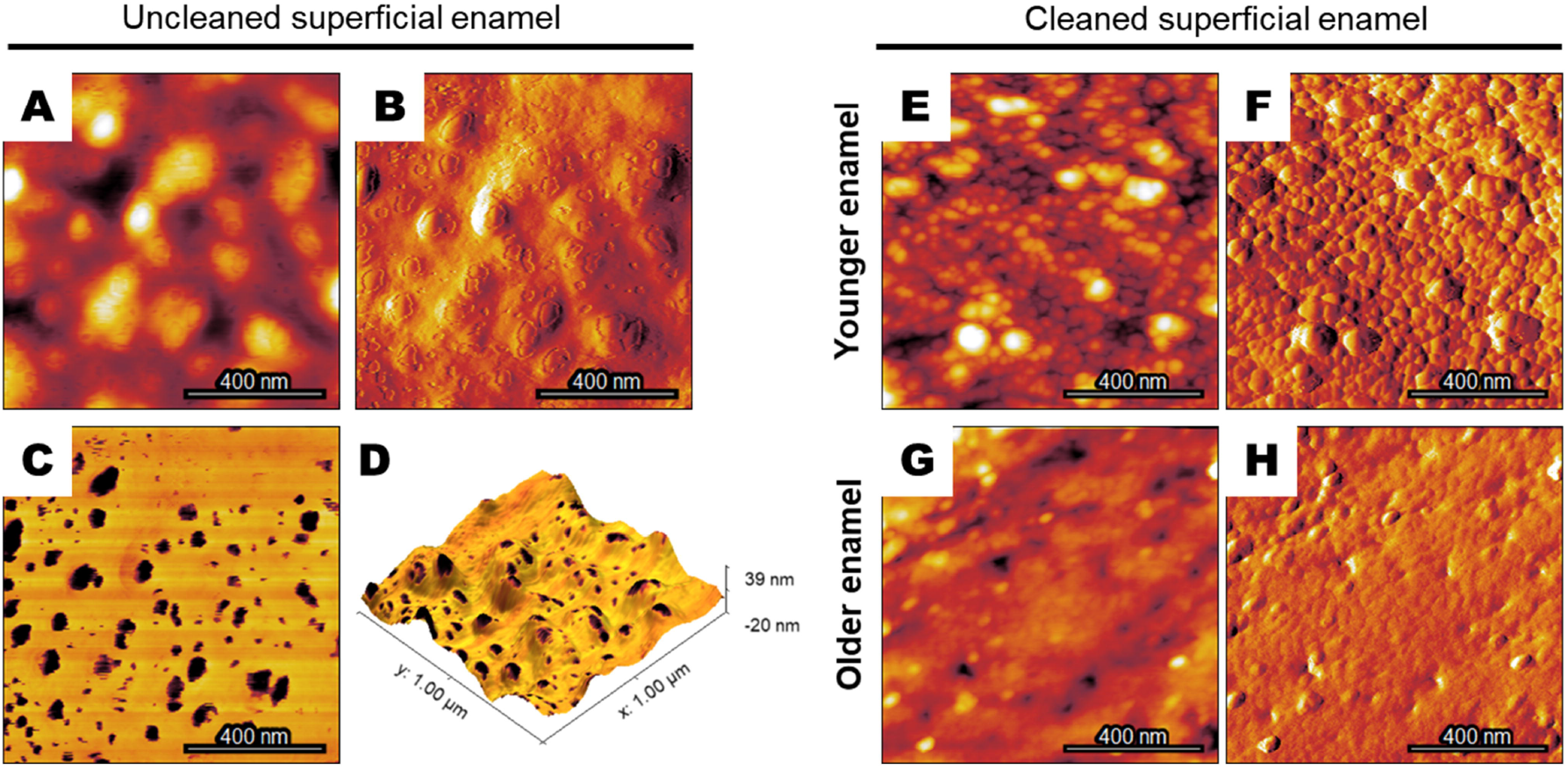
Ultrastructure of superficial dental human enamel. (A) Height, (B) amplitude, (C) phase contrast, and (D) 3D reconstruction images of the superficial enamel layer (in contact with the oral environment), where the presence of a biological pellicle-like structure is observable. (E) Height and (F) amplitude images of cleaned superficial enamel from a younger specimen, showing the characteristic globular morphology associated to the presence of hydroxyapatite crystals. (G) Height and (H) amplitude surfaces in an older sample, displaying a less-defined surface morphology.

After pellicle removal by surface cleaning with dH_2_O and N_2_ airflow, the morphology of enamel was clearly observed in the form of HA crystals, which displayed a globular aspect in high-magnification AFM images (**Figure 1**). Furthermore, enamel in the younger specimen presented clearly demarcated HA crystals on the surface (**Figures 1E and 1F**), while the older specimen appeared to have less defined HA crystals with fewer variations in surface morphology as observed in height and amplitude images (**Figures 1G and 1H**). This observation could potentially be explained by the progressive changes that enamel undergoes during life due to chemical, bacterial, or mechanical factors that can alter its surface morphology^22,23^. Therefore, as enamel is a tissue that once deposited cannot be remodelled or regenerated, the cumulative effect of damage over the years results in surface modifications that can be observed with diverse microscopy approaches^24^. As such, the observation that older enamel is smoother and has less defined HA crystals is expected; but nevertheless, the examination of enamel samples across a wider range of individuals is necessary in the future in order to determine the reproducibility of these observations at a larger population-based level.

### 3.2. Ultrastructure of enamel from tooth cross-sections

As a second step, tooth specimens were sectioned into 200 µm slices and demineralised with a solution of 37% phosphoric acid for 30sec. This demineralisation process is customary during the restorative dental treatment when utilising composite resin-based materials, in order to expose fresh HA crystals on the surface and increase surface energy and micro-retention^25,26^. This process is crucial for the correct adhesion of the restorative material to the enamel surface and long-term restoration survival. Therefore, the resulting sections utilised in this study are representative of clinical enamel surface preparation found in the everyday restorative dentistry setting.

For an initial microscale characterisation, SEM imaging was employed that confirmed etching of enamel by HA removal and showed that different etching morphologies and crystal orientations can be present within one sample (**Figure 2A-D**, obtained from a younger specimen). Correspondingly, when HA crystals are sectioned across their width, they are observed in a globular fashion under AFM similar to the images observed for superficial sound enamel (**Figure 2E and 2F**). Obtaining surface profiles from AFM height images confirms the globular shape and overall dimensions of HA crystals, with a representative size of <80nm (**Figure 2G and 2H**). On the other hand, when crystals were sectioned and exposed longitudinally after acid etching, they were observed in a rod-like fashion under the AFM with lengths <200 nm, consistent with the expected length for individual HA crystals reported previously in the literature (**Figure 2I-L**)^27^. However, it is known that enamel crystals can reach much higher lengths according to their location within the tissue^8,27^. Nevertheless, the characterisation of HA crystal length was not within the scope of this particular work and therefore, only regions with HA crystals observed in globular appearance (such as shown in **Figure 2E**) were utilised for morphological quantification as discussed in the following section.

**Figure 2:**
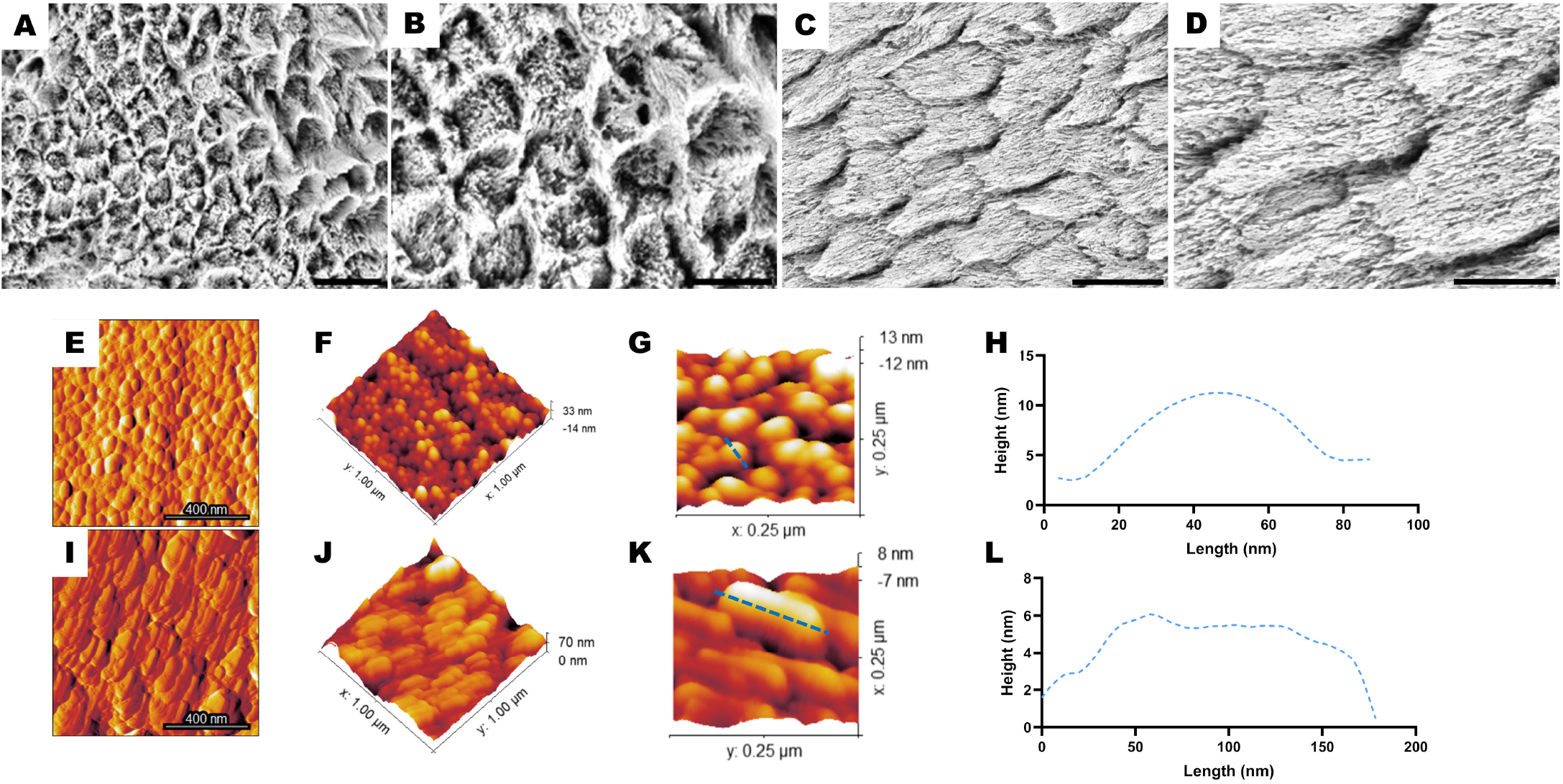
Ultrastructure of hydroxyapatite crystals within human enamel slices following phosphoric acid etching. While in (A) and (B) mineral is mostly removed from the center of enamel rods, (C) and (D) display removal from mostly the interrod area (scale bars A, C: 10µm; B, D: 5µm). Thus, apatite crystals can be exposed either transversally or longitudinally according to their orientation for AFM visualisation. (E) Amplitude and (F) 3D height reconstruction of HA crystals exposed by acid etching (1×1 µm scans) in transversal oriantation. (G) 3D reconstruction of a 250×250nm area of enamel, confirming the globular aspect of HA crystals. (H) Surface profile obtained atop an HA crystal confirming the globular morphology. (I) Amplitude and (J) 3D height reconstruction of HA crystals exposed longitudinally during sample preparation (1×1 µm scans). (K) 3D reconstruction of a 250×250 nm area of enamel, on which the (L) surface profile on a single transversal HA crystal was obtained.

### 3.3. Quantification of HA crystal size and enamel nanoroughness

As a final step, AFM height images from all specimens were utilised to characterise the morphology of HA crystals across all individuals. In order to explore potential anatomical differences within enamel, associated to depth, data was obtained and analysed for both the external and internal areas of the tissue (as shown in **Figure 3A**). Overall, the mean HA width was found to mostly cluster between ∼40-60 nm across all studied groups when the data is pooled from all specimens (**Figure 3B**). These results are consistent with early explorations by Habelitz et al. that also found crystals with a diameter of about 50nm in ground-and-polished enamel specimens^28^. When looking at the group pooled data, we observed a reduction in the diameter of HA crystals for the older specimens compared to younger enamel. Regarding this finding, Zheng et al. observed a reduction in the diameter of HA crystals from worn human enamel surfaces compared to control enamel^19^. Therefore, our observations may be representative of the increased wear expected for older teeth that have been bearing complex force loads within the oral cavity for decades. This is partially believed to be a result of the breaking of HA crystals into smaller ones due to mechanical action as a means to dissipate forces and avoid fracture propagation towards deeper areas of the tooth^29^. Therefore, the reduction of HA crystal width would be expected to occur mostly in the external region of enamel, as shown by our findings. Interestingly, individual variations can be seen within each sample group, particularly among specimens obtained from older tooth samples that display a higher heterogenicity across specimens from different individuals at both the external and internal enamel locations (**Figure 3C**).

**Figure 3:**
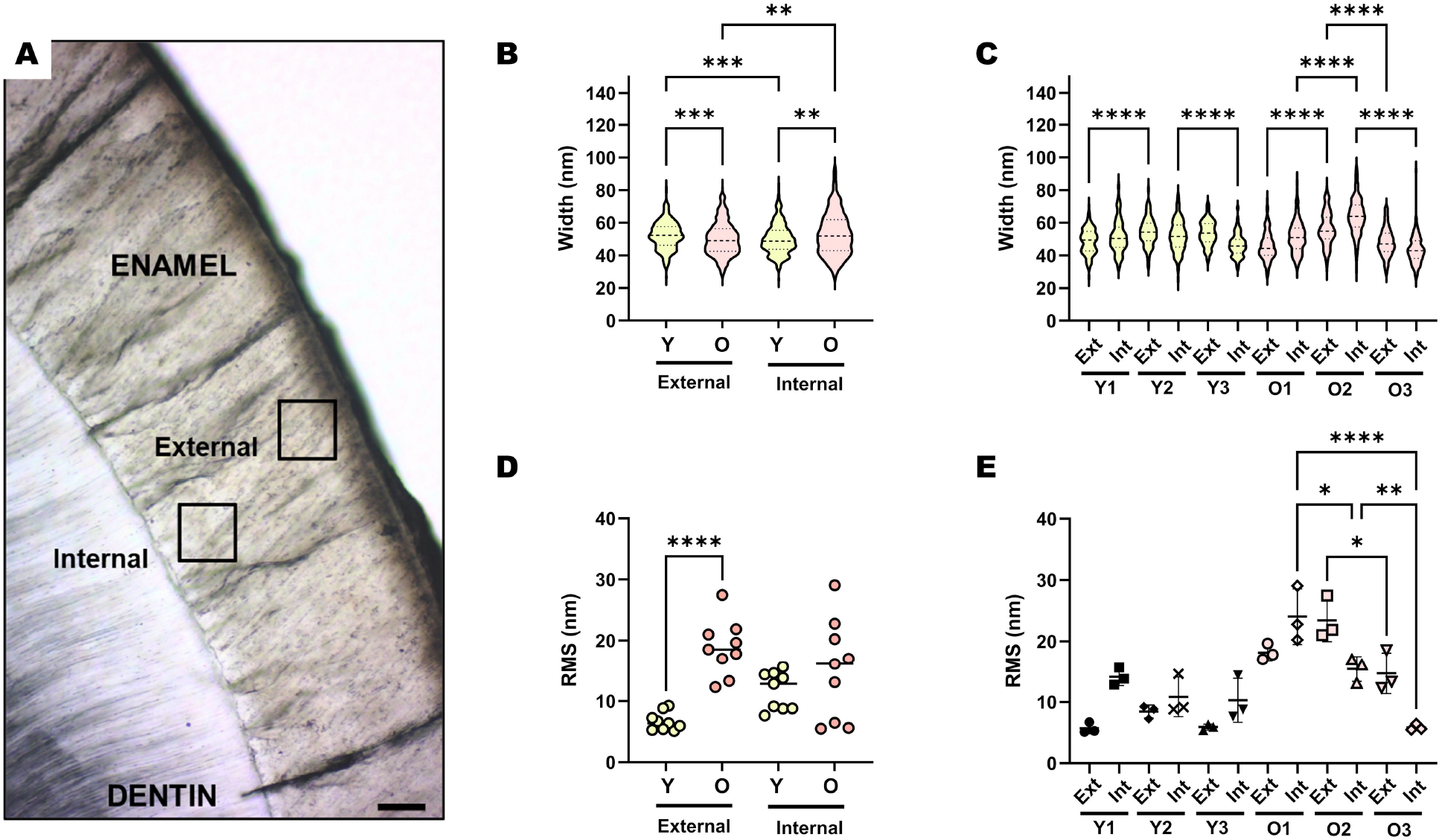
Exploring age-associated quantitative changes in human enamel apatite crystals with AFM. (A) Light microscopy image of a tooth specimen showing the anatomical association between enamel and the underlying dentin layer (scale bar: 100µm). Analysis of enamel was performed at both the external (subsurface) as well as at the internal (vicinity of dentin) areas, as illustrated by the black boxes. (B) Pooled and (C) individual violin plots for HA crystal width at both external and internal locations of enamel for younger (Y) and older (O) specimens. (D) Pooled and (E) individual specimen violin plots for surface roughness (RMS) at both the external and internal locations of enamel (*p<0.05; **p<0.01; ***p<0.001; ****p<0.0001; one-way ANOVA with Tukey’s post-hoc).

Subsequently, the surface roughness of enamel from different specimens was determined with nanoprofilometry derived from AFM height scans. Overall, the RMS values for enamel ranged between ∼5-30 nm across all analysed samples, which are consistent with previous reports in the literature. For example, Lechner et al. reported RMS values of <20 nm for control human incisor enamel samples before demineralisation^30^ and Quartarone et al. reported the average RMS roughness for human enamel to be 50nm^31^. In the present work, we observed a higher surface roughness for the external enamel region in older specimens compared to the younger ones (**Figure 3D**). It has previously been discussed that the increase of enamel surface roughness can be a result of erosive processes due to the consumption of acidic foods and beverages^30^. Therefore, it remains possible that these age-associated changes are a result of chronic exposure of teeth to erosion over time. This is particularly strengthened by the fact that significant differences in roughness were only observed in external enamel, immediately adjacent to the surface of the tooth, and are not observable in the deeper regions of enamel (**Figure 3D**). Also, younger teeth showed remarkable similarity for their surface roughness across all three individual samples (**Figure 3E**). The opposite was observed for older specimens, where important differences were found among individual samples, particularly associated to the internal enamel region that displayed a decrease in surface roughness with increasing ages across the three analysed specimens (**Figure 3E**). As this behaviour is observed only in the internal enamel region, it is not believed to be strongly associated to external factors as is the case with sub-superficial enamel. However, the reduced number of samples (n=3) makes it difficult to determine if this pattern is coincidental, and further work should employ an increased number of specimens across multiple age-groups in order to clarify this observation. Nevertheless, these results confirm the important differences that exist among individuals and biological tissues in *ex-vivo* experiments, and highlight the multifactorial nature of ageing across different individuals.

Overall, this work was an initial exploratory study – with a limited sample size - to determine potential differences in enamel topography, HA crystal size, and surface roughness at the nanoscale associated with ageing. Furthermore, the optimisation of AFM-based approaches for the characterisation of nanoscale qualitative and quantitative changes in enamel is crucial towards future studies with higher sample sizes across populations. In this context, AFM has proven to be a powerful tool for the reproducible ultrastructural characterisation of enamel from human dental specimens with nanoscale precision. Furthermore, the possibility of obtaining a range of qualitative and quantitative information from each surface scan (i.e., topographical characterisation, surface roughness, and HA crystal dimensions) allows for a multiparametric analysis of dental enamel across a range of individuals and specimens with no additional sample preparation. This can preserve the native properties of tissues compared to other microscopy-based approaches and allows a better translation of lab-based results into the clinics.

In this report, our AFM-based technique was utilised to compare different regions of enamel across a small number of individuals from two different age groups, suggesting that older enamel may display particular ageing markers that could play an important role – for example - in restorative dentistry approaches. However, it is important to consider that changes in dental tissues associated to ageing are multifactorial, and may involve several molecular, mechanical, and biological alterations over time^32^. Furthermore, individual differences must be considered when analysing the resulting data, particularly as the process of ageing is not uniform across individuals and may have important differences from a person-to-person basis. Therefore, future work should focus on exploring the ageing of mineralised dental tissues – including enamel - across larger sample sizes in order to compensate for individual differences as well as other potential confounding factors such as behavioural habits and mechanical forces, among others.

## 4. Conclusion

AFM was an effective approach for the characterisation and quantification of human dental enamel ultrastructure at high resolution (nanometre range), in a non-destructive manner and with minimal sample preparation. Despite individual variations in the morphology, size of HA crystals, and surface roughness across samples, older enamel specimens displayed and overall reduced HA size and increased surface roughness in the sub-surface (external) region of the tissue compared to younger enamel. This work further confirms the need to consider individual variations when characterising and quantifying age-associated changes in tissues including mineralised dental tissues such as enamel.

## Data Availability

All data produced in the present study are available upon reasonable request to the authors.

## 5. Acknowledgments

This work was supported by the ANID FONDECYT Iniciación Grant #11180101 and Millennium Science Initiative #P10-035F. The authors would also like to thank Camila Ramos and Dr. Gonzalo Narea for their support with tooth sample collection.

## References

1. Lacruz, R. S., Habelitz, S., Wright, J. T. & Paine, M. L. (2017) Dental enamel formation and implications for oral health and disease. Physiological reviews 97, 939–993.

2. Simmer, J. P. & Fincham, A. G. (1995) Molecular Mechanisms of Dental Enamel Formation. Critical Reviews in Oral Biology & Medicine 6, 84–108.

3. Bartlett, J. D. et al. (2006) Protein–Protein Interactions of the Developing Enamel Matrix. in Current Topics in Developmental Biology vol. 74 57–115 (Academic Press, 2006).

4. Bidlack, F. B., Huynh, C., Marshman, J. & Goetze, B. (2014) Helium ion microscopy of enamel crystallites and extracellular tooth enamel matrix. Frontiers in Physiology 5, 395.

5. Besnard, C. et al. (2021) Analysis of in vitro demineralised human enamel using multi-scale correlative optical and scanning electron microscopy, and high-resolution synchrotron wide-angle X-ray scattering. Materials & Design 206, 109739.

6. Lubarsky, G. V, D’Sa, R. A., Deb, S., Meenan, B. J. & Lemoine, P. (2012) The role of enamel proteins in protecting mature human enamel against acidic environments: a double layer force spectroscopy study. Biointerphases 7, 14.

7. Daculsi, G. & Kerebel, B. (1978) High-resolution electron microscope study of human enamel crystallites: Size, shape, and growth. Journal of Ultrastructure Research 65, 163–172.

8. Beniash, E. et al. (2019) The hidden structure of human enamel. Nature Communications 10, 4383.

9. Sato, T., Takagaki, T., Hatayama, T., Nikaido, T. & Tagami, J. (2021) Update on Enamel Bonding Strategies . Frontiers in Dental Medicine vol. 2.

10. Hillmann, G. & Geurtsen, W. (1997) Light-microscopical investigation of the distribution of extracellular matrix molecules and calcifications in human dental pulps of various ages. Cell and Tissue Research 289, 145–154.

11. Shinno, Y. et al. (2016) Comprehensive analyses of how tubule occlusion and advanced glycation end-products diminish strength of aged dentin. Scientific Reports 6,.

12. Gautier, H. O. B. et al. (2015) Chapter 12 - Atomic force microscopy-based force measurements on animal cells and tissues. in Methods in Cell Biology (ed. Paluch, E. K.) vol. 125 211–235 (Academic Press, 2015).

13. Stylianou, A., Kontomaris, S.-V., Grant, C. & Alexandratou, E. (2019) Atomic Force Microscopy on Biological Materials Related to Pathological Conditions. Scanning 2019, 8452851.

14. Ando, T., Uchihashi, T. & Kodera, N. (2013) High-speed AFM and applications to biomolecular systems. Annual review of biophysics 42, 393–414.

15. Aguayo, S. & Bozec, L. (2016) Mechanics of bacterial cells and initial surface colonisation. Advances in Experimental Medicine and Biology vol. 915 (2016).

16. Torres-Gallegos, I. et al. (2012) Enamel roughness and depth profile after phosphoric acid etching of healthy and fluorotic enamel. Australian Dental Journal 57, 151–156.

17. Poggio, C., Ceci, M., Beltrami, R., Lombardini, M. & Colombo, M. (2014) Atomic force microscopy study of enamel remineralization. Annali di stomatologia 5, 98–102.

18. Meredith, L., Farella, M., Lowrey, S., Cannon, R. D. & Mei, L. (2017) Atomic force microscopy analysis of enamel nanotopography after interproximal reduction. American Journal of Orthodontics and Dentofacial Orthopedics 151, 750–757.

19. Zheng, J. et al. (2013) Microtribological behaviour of human tooth enamel and artificial hydroxyapatite. Tribology International 63, 177–185.

20. Chawhuaveang, D. D. et al. (2021) Acquired salivary pellicle and oral diseases: A literature review. Journal of Dental Sciences 16, 523–529.

21. Siqueira, W. L., Custodio, W. & McDonald, E. E. (2012) New Insights into the Composition and Functions of the Acquired Enamel Pellicle. Journal of Dental Research 91, 1110–1118.

22. Donovan, T., Nguyen-Ngoc, C., Abd Alraheam, I. & Irusa, K. (2021) Contemporary diagnosis and management of dental erosion. Journal of Esthetic and Restorative Dentistry 33, 78–87.

23. Pitts, N. B. et al. (2017) Dental caries. Nature Reviews Disease Primers 3,.

24. Xiao, H. et al. (2021) Protective effects of two food hydrocolloids on dental erosion: Nanomechanical properties and microtribological behavior study. Friction 9, 356–366.

25. Bertacci, A., Lucchese, A., Taddei, P., Gherlone, E. F. & Chersoni, S. (2014) Enamel Structural Changes induced by Hydrochloric and Phosphoric Acid Treatment. Journal of Applied Biomaterials & Functional Materials 12, 240–247.

26. Zhu, J. J., Tang, A. T. H., Matinlinna, J. P. & Hägg, U. (2014) Acid etching of human enamel in clinical applications: A systematic review. The Journal of Prosthetic Dentistry 112, 122–135.

27. Daculsi, G., Menanteau, J., Kerebel, L. M. & Mitre, D. (1984) Length and shape of enamel crystals. Calcified Tissue International 36, 550–555.

28. Habelitz, S., Marshall, S. J., Marshall, G. W. & Balooch, M. (2001) Mechanical properties of human dental enamel on the nanometre scale. Archives of Oral Biology 46, 173–183.

29. Zheng, S. Y. et al. (2011) Investigation on the microtribological behaviour of human tooth enamel by nanoscratch. Wear 271, 2290–2296.

30. Lechner, B.-D., Röper, S., Messerschmidt, J., Blume, A. & Magerle, R. (2015) Monitoring Demineralization and Subsequent Remineralization of Human Teeth at the Dentin–Enamel Junction with Atomic Force Microscopy. ACS Applied Materials & Interfaces 7, 18937–18943.

31. Quartarone, E., Mustarelli, P., Poggio, C. & Lombardini, M. (2008) Surface kinetic roughening caused by dental erosion: An atomic force microscopy study. Journal of Applied Physics 103, 104702.

32. Schuh, C. M. A. P. et al. (2022) Nanomechanical and Molecular Characterization of Aging in Dentinal Collagen. Journal of Dental Research 00220345211072484 doi:10.1177/00220345211072484.

